# Lifespan investigation of brain volumetric changes associated with substance use disorders

**DOI:** 10.1101/2025.05.28.25328476

**Authors:** Runye Shi, Shitong Xiang, Dag Alnæs, Di Chen, Zheng Chen, Tobias Banaschewski, Gareth J. Barker, Arun L.W. Bokde, Sylvane Desrivières, Herta Flor, Hugh Garavan, Penny Gowland, Antoine Grigis, Andreas Heinz, Jean-Luc Martinot, Marie-Laure Paillère Martinot, Eric Artiges, Frauke Nees, Dimitri Papadopoulos Orfanos, Luise Poustka, Michael N. Smolka, Sarah Hohmann, Nilakshi Vaidya, Henrik Walter, Robert Whelan, Gunter Schumann, Barbara J. Sahakian, Lars T. Westlye, Trevor W. Robbins, Xiaolei Lin, Tianye Jia, Jianfeng Feng, IMAGEN Consortium

## Abstract

Substance use disorder (SUD) stands as a critical public health concern, contributing to substantial morbidity, mortality and societal costs. The effects of SUD on structural brain changes have been well documented. However, the neural mechanisms underlying SUD and the spatial-temporal volumetric changes associated with SUD remained underexplored. In this investigation, neuroimaging, behavioral and genomic data across four large population cohorts jointly covering the full lifespan were harmonized, and whole-brain volumetric trajectories between substance use disorders (SUDs) and healthy controls (HCs) were compared, revealing the potential neurobiological mechanisms and the genomic basis underlying SUD. Results highlighted three distinct life stages critical for the development of SUD: 1) adolescence to early adulthood (before 25y), where SUD is suspected to be the consequence of prefrontal-subcortical imbalance during neurodevelopment; 2) early-to-mid adulthood (25y – 45y), where SUD was strongly associated with compulsivity-related brain volumetric changes; 3) mid-to-late adulthood (after 45y), where SUD-related brain structural changes could be explained by neurotoxicity. Results were externally validated both via longitudinal analysis of these population cohorts and in independent cross-sectional samples. In summary, our study demonstrated the lifespan whole-brain volumetric changes associated with SUD, revealed potential neurobehavioral mechanisms for the development of SUD, and suggested critical time window for effective prevention and treatment of SUD.

## Introduction

In recent years, there has been increasing research interest for human lifespan neuroscience, emphasizing the challenges of understanding mental health and human behaviors associated with social and physiological characteristics across different age stages^1^. While most psychiatric disorders onset before early adulthood^2^, possibly partly attributed to puberty-related hormonal changes and unbalanced brain development^3^, they demonstrated distinct lifetime prevalence and longitudinal trajectories. Given these complexities, understanding whether the neural signatures and behavioral profiles of different neuropsychiatric disorders follow distinct developmental trajectories or represent a homogeneous continuum remains an open question. Substance use disorder (SUD), which often emerges during adolescence and persists across the entire lifespan, provides a unique case for investigating the lifespan trajectories associated with brain volumetric changes and neurocognitive performances.

SUD is a complex health condition characterized by compulsive, persistent and risky substance use or misuse behaviors in inappropriate situations^4^, exerting a lifespan impact on both mental and physical health^5–7^, including cognitive impairments, behavioral dysfunctions and chronic illnesses such as cardiovascular disease and diabetes^8–10^. The high prevalence of SUD further amplifies its negative impacts. In 2022, an estimated 17.3% (48.7 million) of the population aged 12 years or older in the United States had SUD according to a self-reported national survey of approximately 70 thousand participants^11^, including 29.5 million having an alcohol use disorder, 27.2 million having drug use disorder, and 8.0 million having both. In Europe, an estimated 29% (83.4 million) of those aged 15 years or older reported previous use of illicit drugs^12^. Some risk factors may exert different impacts at different life stages, with higher BMI positively correlated with alcohol consumption in adolescent girls^13,14^ and negatively associated with past-year alcohol abuse risk in adult women^15,16^. Other factors were characterized by time-specific effects, for example, the shift from an enriched to standard environment in mice was only found to enhance the sensitivity of drug reward during early life stages^17^. Further, the progression of neuropsychiatric disorders may vary across time with differential impacts on the brain, likely due to time-varying neuroplasticity and long-term homeostatic compensation mechanisms taking place at different life stages^18,19^. For instance, the risk of substance dependence is reduced by 4%–5% for each year of delayed substance use initiation from age 13 to 21 years old when the brain gradually matures^20^, while significantly increased in older life stages with reduced drug metabolism and aging^21,22^. Findings on longitudinal SUD-related changes in brain regions were inconsistent across studies, with some studies focusing on SUDs among adolescents, while others specifically investigating early and mid-to-late adulthood^23–27^. Despite these insights, a comprehensive lifespan approach on the neural mechanisms underlying SUD remains underexplored.

To address these questions, we utilized data from the Adolescent Brain Cognitive Development (ABCD) study^28^, the IMAGEN study^29^, the Human Connectome Project (HCP)^30^ and UK Biobank (UKB)^31^ with participants ranging from adolescence to early adulthood and extending into older ages. While previous studies have revealed that both brain structures and functional connectivity undergo characteristic changes across the lifespan^32–34^, a normative model was introduced to deal with the heterogeneity within population cohorts due to age and other study-specific characteristics, treating mental disorders and specific behaviors as deviations from a normative developmental trajectory^35^. Normative models were constructed for each brain region of interest (ROI) using the Generalized Additive Model for Location, Scale and Shape (GAMLSS)^36^, which have demonstrable accuracy for depicting life-course trajectories of brain morphology^32,37^. Morphological characteristics were quantified as centile scores for all individuals to adjust for potential confounding. Separate volumetric trajectories were constructed for individuals with SUD and healthy controls (HCs). In general, compared to HCs, individuals with SUD had lower grey matter volume (GMV) in cortical regions, with differences following an inverted U-shape over time, while the GMV differences in subcortical regions gradually decreased over time. Both volumetric and neurobehavioral results were successfully validated using Enhanced Nathan Kline Institute-Rockland Sample (NKI-RS), Cambridge Centre for Ageing and Neuroscience (Cam-CAN), and Brains and Minds in Transition (BRAINMINT) samples. Correlations between GMV centiles and neurobehavioral scores were investigated throughout the life span using common factors and pooled meta-analysis of effect sizes due to inconsistencies in measurements across studies. Finally, to understand the genomic basis underlying the lifespan SUD trajectories, genome-wide association study (GWAS) and genomic correlation analysis were performed, and genetic variants associated with GMV-predicted SUD were identified using conjunctional FDR^38^ (conjFDR).

## Results

### Definition of SUD across the lifespan

A total of 51,467 participants aged 9y – 70y with 53,199 neuroimaging scans across four cohorts (ABCD, IMAGEN, HCP and UKB) were included in the discovery set, and a total of 2,127 participants aged 12y – 89y from the cross-sectional NKI-RS, Cam-CAN and BRAINMINT studies were included for validation. Due to different accessibility of addictive substances used between adolescents and adults, an adaptive definition of SUD (among alcohol, tobacco, marijuana and any other reported addictive substances) was employed: for adolescents, those engaging in any addictive substances intake behavior or meeting the criteria of child addiction scale were classified as SUD; for adults, those meeting the clinical criteria of substance dependence or ranking in the top 25% of substance use frequency were classified as SUD. Meanwhile, participants with no substance use at any time throughout the life span or occasional substance use during adulthood were considered as HCs. Detailed descriptions of the definitions of SUD and HC were provided in the Methods section. Based on the criteria above, a total of 6,992 and 711 participants were identified as SUDs, while 18,659 and 1,416 were included as HCs in the discovery and validation sets, respectively. Overall, the proportion of SUDs among the study population first increased with age, peaked at approximately the age of 25y, and then started to decrease thereafter, as well as the overlap of participants across different substance use groups. (Supplementary Fig. 1).

Cross-sectional investigation (Supplementary Table 1) indicated consistent differences between SUDs and HCs on demographic characteristics across the lifespan: SUDs are more likely to be males (*X*^2^ = 8.80, *P =* 0.003 for ABCD; *X*^2^ = 5.29, *P =* 0.023 for IMAGEN-BL; *X*^2^ = 39.53, *P* < 0.001 for IMAGEN-FU2; *X*^2^ = 39.03, *P* < 0.001 for IMAGEN-FU3; *X*^2^ = 41.71, *P* < 0.001 for HCP; *X*^2^ = 1248.3, *P* < 0.001 for UKB), having lower intelligence score (*d* = -0.40, *P* < 0.001 for ABCD; *d* = -0.20, *P =* 0.015 for IMAGEN-BL; *d* = -0.04, *P =* 0.031 for UKB), residing in poorer socioeconomic conditions (*d* = -0.48, *P* < 0.001 for ABCD; *d* = -0.16, *P =* 0.046 for IMAGEN-BL; *d* = -0.23, *P =* 0.002 for HCP; *d* = -0.27, *P* < 0.001 for UKB), and having experienced more negative life events (*d* = 0.44, *P* < 0.001 for ABCD; *d* = 0.17, *P =* 0.009 for IMAGEN-FU2; *d* = 0.19, *P =* 0.010 for HCP; *d* = 0.22, *P* < 0.001 for UKB). On the contrary, we observed inconsistent BMI between SUDs and HCs across different life stages: SUDs are more likely to be associated with larger BMI during adolescence, but with lower BMI during late adulthood (*d* = 0.28, *P =* 0.011 for ABCD; *d* = 0.18, *P =* 0.024 for IMAGEN-BL; *d* = 0.15, *P =* 0.025 for IMAGEN-FU2; *d* = -0.05, *P =* 0.005 for UKB).

### Lifespan volumetric brain differences between SUDs and HCs

Twenty-four brain regions of interest (ROIs) involved in the development of SUD were selected as suggested by previous studies^39,40^. These regions (ventral/dorsal striatum, amygdala, hippocampus, insula, anterior cingulate and prefrontal cortex) are key components in the addiction neurocircuitry, and are involved in reward processing, motivation, contextual memory, interoception, and executive function. The GAMLSS model^32^ was employed to harmonize neuroimaging data across the lifespan and construct age-specific normative ranges of HCs for each ROI (Supplementary Fig. 2). Cubic spline model was developed to estimate the age-specific GMV trajectories for SUDs and HCs separately, adjusting for sex, handedness and studies.

Dynamic GMV volumetric differences across the lifespan were observed between SUDs and HCs (Fig. 1). During childhood and adolescence, compared to HCs, SUDs showed higher volumetric GMV in the bilateral putamen and lower GMV in the bilateral nucleus accumbens, insula, frontal pole, orbitofrontal, superior frontal, rostral anterior cingulate (ACC) and left caudal ACC. Notably, participants with SUD experienced delayed development in the left medial orbitofrontal and bilateral nucleus accumbens, regions that may mediate dopamine-related goal-directed behavior^41,42^. This is also consistent with the finding that smaller left ventral mediofrontal cortex was causally associated with smoking initiation among adolescents^26^. However, these GMV volumetric differences observed in early life stages gradually diminished with brain development, which indicated that early brain volumetric differences between SUDs and HCs could possibly contribute to the substance use. Later during adulthood, SUDs exhibited relatively lower GMV and faster GMV decreasing rate in most ROIs. Starting from amygdala, GMV differences between SUDs and HCs became increasingly apparent in the hippocampus, nucleus accumbens, medial orbitofrontal cortex, and right lateral orbitofrontal cortex. involving hippocampus, nucleus accumbens, medial orbitofrontal cortex and right lateral orbitofrontal cortex. Notably, higher volumetric GMV in SUDs was also observed in the salience network (SN; also called mid-cingulo-insular network, including insula, rostral ACC and left lateral orbitofrontal cortex), with significant differences starting at approximately 30y, peaking at mid 40ys and gradually diminishing after 60y. According to the lifespan volumetric differences between SUDs and HCs, ROIs can be subdivided into three categories: those differentiating SUDs and HCs at early stages of life, including bilateral frontal pole, insula, orbitofrontal, superior frontal, rostral ACC, nucleus accumbens and putamen; those differentiating SUDs and HCs at intermediate stages of life, including bilateral insula, lateral orbitofrontal and rostral ACC; and those differentiating SUDs and HCs at later stages of life, including left frontal pole, lateral orbitofrontal, bilateral medial orbitofrontal, superior frontal, amygdala, hippocampus and nucleus accumbens.

**Fig. 1.**
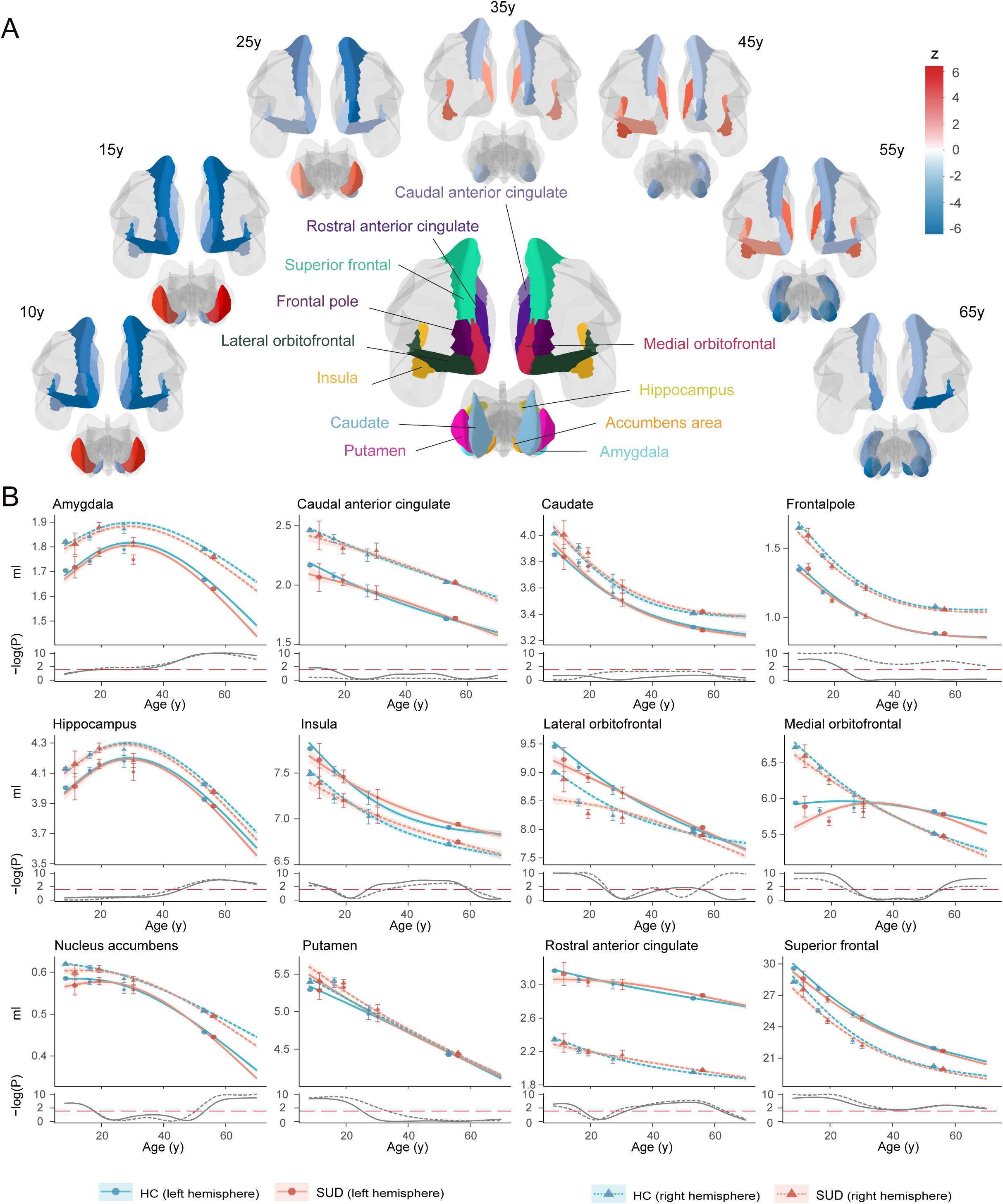
Lifespan GMV trajectories for SUDs and HCs. (A) Visualization of ROIs with significant differences between SUDs and HCs at different life stages throughout the lifespan. Blue indicates lower GMV in SUDs compared to HCs, while red indicates the opposite. (B) Estimated lifespan GMV trajectories for SUDs (red) and HCs (blue) with 95% confidence bands, adjusting for sex, handedness and study. Observed mean GMVs from each study were displayed as points, with errors bars representing standard errors (Top). Two sample t-test was used to compare GMV between SUDs and HCs across the lifespan, with Benjamini-Hochberg correction for multiple testing within each ROI. Log p-values from the tests were reported against the significance level (dashed red line) (Bottom).

The lifespan volumetric patterns between SUDs and HCs were validated via both longitudinal analysis of the neuroimaging data using ABCD and UKB (Supplementary Fig. 3) and cross-sectional analysis of the external NKI-RS, Cam-CAN and BRAINMINT datasets (Supplementary Fig. 4). Due to the limited sample size and relatively large standard errors in validation samples, it was impractical to obtain statistically significant results as in the discovery set. Therefore, the external validation largely relied on observing similar GMV volumetric patterns between SUDs and HCs, with moderate to strong correlations in 71% significant comparative z-statistics (17 out of 24 ROIs; *r* = 0.25-0.95) between the discovery and validation set.

Since the change of SUD definition from adolescence to adulthood may lead to the discontinuity of volumetric GMV changes, a sensitivity analysis was conducted by modifying the SUD definition for adults such that those with above experimental substance use or regular substance use were also included as SUDs. Despite inconsistencies in certain ROIs, the overall lifespan GMV patterns between SUDs and HCs remain similar (Supplementary Fig. 5).

Although the lifespan volumetric patterns between SUDs and HCs were relatively consistent in the left and right hemispheres, we noticed that differences in the right hemisphere were more pronounced in putamen, frontal pole and lateral orbitofrontal regions compared to the left hemisphere, while differences in the left hemisphere were greater in the medial orbitofrontal and caudal ACC. These observed volumetric differences in the left and right hemispheres may suggest potential roles of the corresponding hemisphere during SUD development.

Further, we investigated the lifespan volumetric trajectories for SUDs separately for each addictive substance, including alcohol, tobacco, marijuana and any other type of addictive drugs. The lifespan volumetric differences between SUDs and HCs showed highly similar patterns for alcohol, tobacco and marijuana (Supplementary Fig. 6-8), with slightly higher GMV in insula, rostral ACC and lateral orbitofrontal observed for alcohol and marijuana users during early adulthood. However, inconsistent results were observed for drug users, who showed higher GMV than HCs during late adulthood (Supplementary Fig. 9). This inconsistency was likely due to the small sample size and less robust model fitting of GMV trajectories for drug users.

### Lifespan associational patterns between neurobehavioral performances and SUD

Since SUD is often associated with neurobehavioral changes, we next investigated how this association varies with age across lifespan by comparing SUD-related neurobehavioral performances between SUDs and HCs in each study. Significant neurobehavioral differences in SUD emerge progressively over the life-course (Fig. 2A and Supplementary Tables 2-5). During pre-adulthood (in ABCD and IMAGEN studies), SUDs were more likely to be engaged in rule-breaking behaviors, had more conduct problems and higher impulsivity scores, especially in sensation seeking, based on self-reported questionnaires and cognitive tests such as Monetary Incentive Delay task (MID) in ABCD and Monetary-Choice Test (KIRBY) in IMAGEN. It is noteworthy that SUDs did not show preference for immediate rewards facing large delayed rewards in the KIRBY compared to HCs (*d* = 0.10, *P_adj_* = 0.154), nor did they exhibit differences in reaction time on the Stop Signal Task (SST) (*d* = -0.06, *P_adj_* = 0.363), indicating that their impulsive behaviors were relatively limited. We also found no evidence toward some aspects of impaired executive functioning among SUDs via the Dimensional Change Card Sort Test in ABCD (cognitive flexibility) (*d* = -0.20, *P_adj_* = 0.072) and the spatial working memory test in IMAGEN (*d* = -0.07, *P_adj_* = 0.363), although SUDs exhibited lower fluid (*d* = -0.26, *P_adj_* = 0.028) and crystallized cognitive abilities than HCs (*d* = -0.40, *P_adj_* = 6.51×10^-4^). During early adulthood (in HCP study), SUDs started to show lower self-awareness, as measured by NIH Toolbox meaning and purpose in life (*d* = -0.31, *P_adj_* = 2.95×10^-4^), and life satisfaction (*d* = -0.19, *P_adj_* = 0.032), while continued to have higher rule-breaking and impulsivity scores as measured by questionnaires compared to HCs. They also display steeper delay discounting across all reward sizes, including larger delayed rewards, suggesting potentially increased impulsivity for SUDs when transitioning from adolescence to adulthood. Finally, during late adulthood (in UKB), SUDs exhibited both increased mood problems and worse neurocognitive performances for executive functions compared to HCs. The lifespan associational patterns between neurobehavioral performances and SUDs were partially validated using the NKI-RS study with a broader age range (Supplementary Fig. 10), where we observed consistently higher rule-breaking and risk-taking scores for SUDs compared to HCs during both pre-adulthood and adulthood, with effect sizes decreasing over time.

**Fig. 2.**
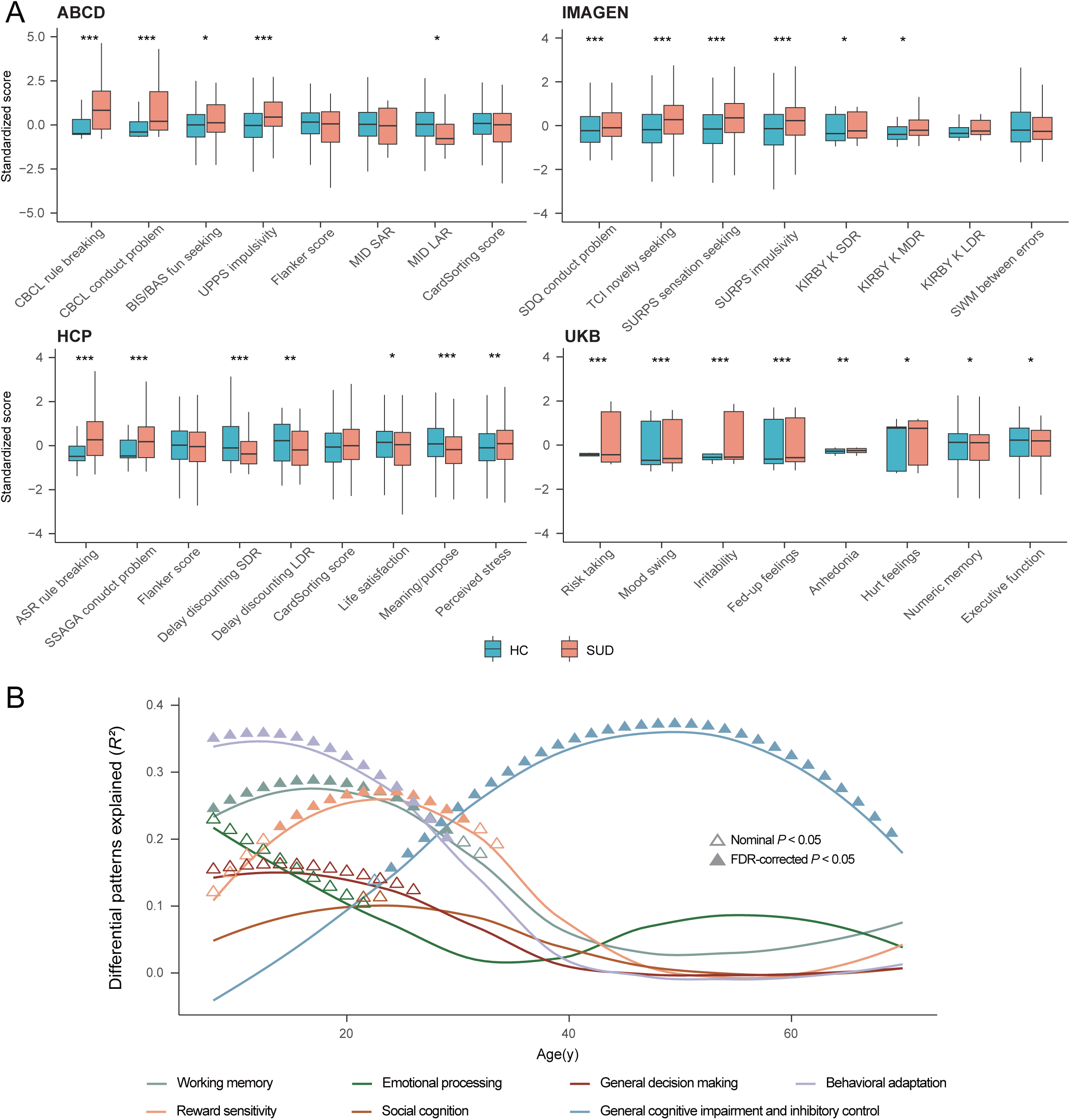
Comparison of behavioral and neurocognitive performances between SUDs and HCs. (A) Comparison of behavioral and neurocognitive performances between SUDs and HCs for all study cohorts, adjusting for sex, handedness and age. Two-tailed two-sample t-test was used, with BH-FDR correction for multiple testing within studies. CBCL, Child Behavior Checklist; BIS/BAS, Behavioral Activation and Behavioral Inhibition Scales; UPPS, Impulsive behavior scale; Flanker, Flanker Inhibitory Control and Attention Test; MID Monetary Incentive Delay task; S/LAR, small/large anticipated reward; SDQ, Strengths and Difficulties Questionnaire; TCI, Temperament and Character Inventory; SURPS, Substance Use Risk Profile Scale; KIRBY, Monetary-Choice Questionnaire; S/M/LDR, small/median/large delayed reward; ASR, Achenbach Adult Self Report; SSAGA, Semi-Structured Assessment for the Genetics of Alcoholism. *** *P_adj_*<0.001; ** *P_adj_*<0.01; * *P_adj_*<0.05. (B) Volumetric differences between SUDs and HCs that can be explained by cognitive factors. Seven factors were extracted from 133 cognitive terms, and were named based on their first ten cognitive terms. Univariate linear regression was conducted between factor-level cognitive brain atlas and GMV differential patterns at each 0.5-year time interval. Triangles indicate positive-sided significance based on spin permutations of brain atlases, with BH-FDR method used for multiple testing. ▲: BH-FDR-corrected *P*<0.05; Δ: nominal *P*<0.05.

Continuous SUD-related cognitive performances were derived from publicly available cognitive brain atlases^43^ across the lifespan, by correlating factor-level cognitive brain atlas with volumetric brain differences between SUDs and HCs. Seven factors (working memory, emotion processing, reward sensitivity, social cognition, general decision making, general cognitive impairment and inhibitory control, and behavioral adaptation) were extracted from 133 cognitive terms, which explained 91% of the total variances (Supplementary Table 6, Supplementary Fig. 11-12). Working memory, behavioral adaptation, and reward sensitivity were primarily associated with SUD during early life stages. Specifically, the prominent association between reward sensitivity and SUD in early adulthood aligned with our analytical results using IMAGEN and HCP. In contrast, the general cognitive impairment and inhibitory control – SUD association kept increasing and reached its peak during mid-to-late adulthood. This observed shift in the cognitive profiles linked to SUD across the lifespan is consistent with the from-impulsivity-to-compulsivity addiction hypothesis^41,44^.

Nonetheless, it should be noted that despite progressively greater impulsivity found among SUDs in terms of fun seeking, sensation seeking and delay discounting, no significant differences were found between SUDs and HCs in terms of the Flanker inhibitory control and attention test. This suggests that when generalizing conclusions, one needs to consider issues such as different functionality of the composite dimension of impulsivity^45^.

Having observed the lifespan associational patterns between neurobehavioral performances and SUDs, we next explored the underlying neurobiological mechanisms via lifespan trajectories of volumetric GMV (Fig. 3). Generalized rule-breaking and impulsivity scores were calculated by factor analysis to accommodate for measurement variability across studies (Supplementary Fig. 13). Correlational analyses between neurobehavioral performances and volumetric GMV were jointly conducted in pre-adulthood for ABCD and IMAGEN, while separately in adulthood for HCP. Consistent with the volumetric patterns during pre-adulthood, higher impulsivity was associated with higher volumetric GMV in dorsal striatum (bilateral caudate and right putamen) and lower GMV in prefrontal (bilateral lateral orbitofrontal, frontal pole and left medial orbitofrontal)/rostral ACC. Later in adulthood, higher impulsivity scores were associated with lower GMV in bilateral caudate, frontal pole, lateral orbitofrontal, rostral ACC, right nucleus accumbens and left medial orbitofrontal cortex. In contrast, significant negative correlation was observed between rule-breaking scores and GMV in rostral/dorsal striatum, prefrontal regions, and ACC during pre-adulthood, with no significant correlations observed in adulthood. These results were robust when adjusting for rule-breaking scores between impulsivity and brain regions, and vice versa (Supplementary Fig. 14).

**Fig. 3.**
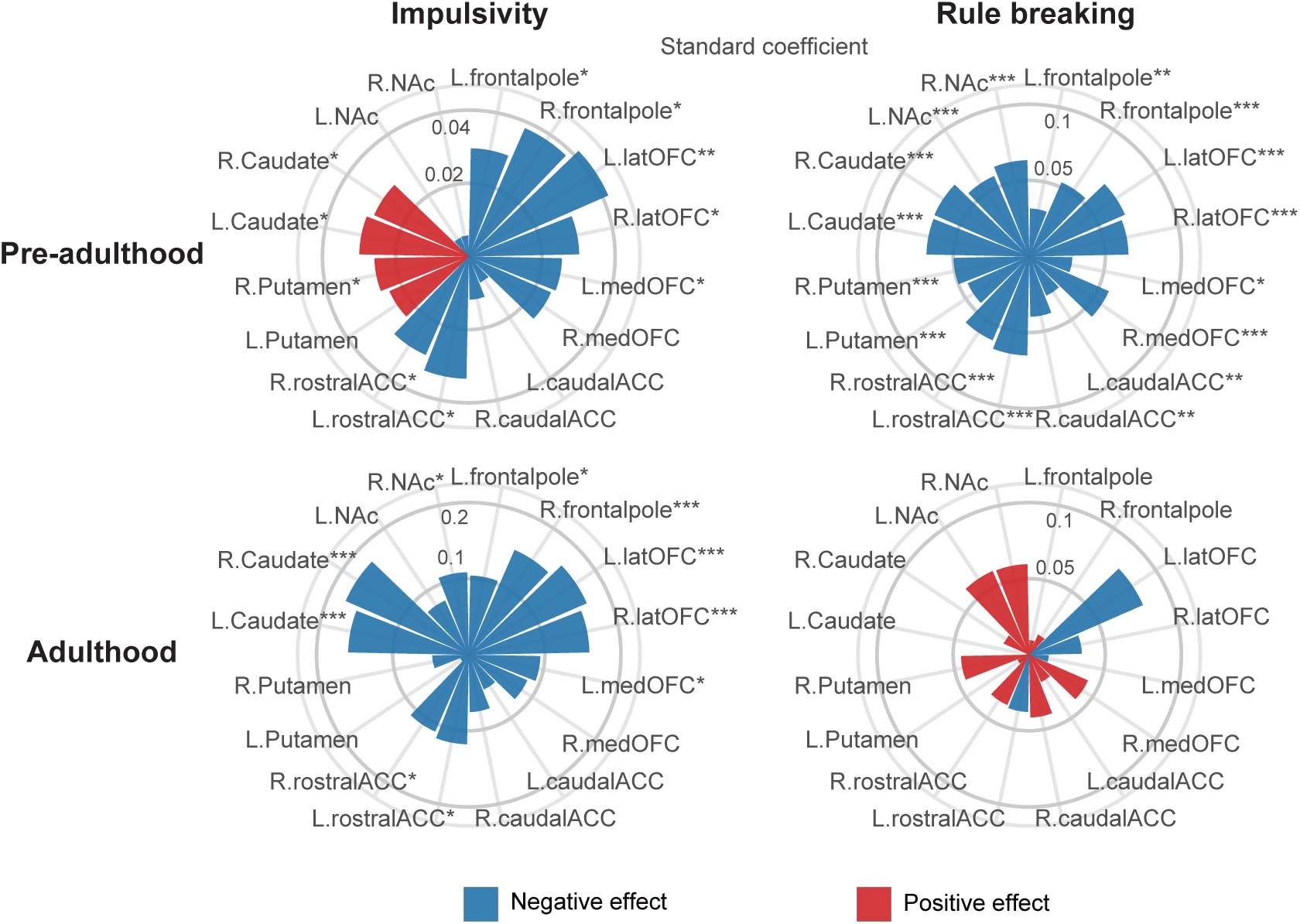
Correlation between impulsivity/rule breaking scores and ROI centiles during pre-adulthood and adulthood. Distinct correlation patterns were observed in pre-adulthood (in ABCD and IMAGEN) and adulthood (in HCP). Impulsivity and rule breaking scores were obtained from factor anlaysis. Two-tailed t-test was used and BH-FDR method was used for multiple testing. latOFC, lateral orbitofrontal; medOFC, medial orbitofrontal; ACC, anterior caudal cingulate; NAc, nucleus accumbens.

### Longitudinal investigation of adolescent SUD development

While cross-sectional lifespan studies have provided valuable insights into SUD, understanding the early factors that contribute to substance use initiation in adolescence from a longitudinal perspective is also crucial for elucidating the mechanisms of SUD development in adolescence and informing prevention strategies. Thus, we investigate whether pre-adulthood volumetric differences could contribute to the initiation of substance use, along with any behavioral evidence. To test this, pre-adulthood HCs at baseline were divided into two subgroups, with the first subgroup developing SUDs in the follow-up visits (hereafter called follow-up SUDs) and the other subgroup remaining as HCs (hereafter called follow-up HCs). 1,930 out of 11,085 (17.41%) pre-adulthood HCs in ABCD and 549 out of 987 (55.62%) pre-adulthood HCs at baseline in IMAGEN were identified as follow-up SUDs, respectively. In ABCD, follow-up SUDs tend to have higher GMV in bilateral caudate (left: *d* = 0.05, *P_adj_* = 0.031; right: *d* = 0.06, *P_adj_* = 0.018) and putamen (left: *d* = 0.05, *P_adj_* = 0.028; right: *d* = 0.06, *P_adj_* = 0.018), and lower GMV in left medial orbitofrontal (*d* = -0.06, *P_adj_* < 0.001), right lateral orbitofrontal (*d* = -0.02, *P_adj_* = 0.045), right frontal pole (*d* = -0.03, *P_adj_* = 0.046), bilateral rostral ACC (left: *d* = -0.05, *P_adj_* = 0.009; right: *d* = -0.05, *P_adj_* = 0.018) and caudal ACC (left: *d* = -0.04, *P_adj_* = 0.009; right: *d* = -0.04, *P_adj_* = 0.009 (Supplementary Fig. 15). These results were consistent with the volumetric differences between SUDs and HCs observed in the early-life stages, and indicated an important role of the prefrontal-striatum imbalance in the onset of SUD. Due to smaller sample size and potentially improvement of the prefrontal-striatum imbalance among adolescents, only lower volumetric GMV in medial orbitofrontal was observed in the follow-up SUDs in IMAGEN (Supplementary Fig. 10). Additionally, we examined whether resilience to pre-adulthood prefrontal-subcortical imbalance may depend on the severity of SUD, and found no significant correlations between longitudinal GMV changes and quantities of substance used or levels of substance dependency (Supplementary Table 7). Regarding behavioral performance comparisons, follow-up SUDs exhibited higher total impulsivity score and greater sensation seeking behaviors as measured by behavioral questionnaires in both ABCD and IMAGEN, whereas they also showed more conduct problems and rule-breaking tendencies in IMAGEN (Supplementary Table 8). However, no differences in rule breaking behavior were observed between follow-up SUDs and follow-up HCs in ABCD.

### SUD-related genomics

To elucidate the genomic basis underlying the lifespan volumetric and neurobehavioral differences between SUDs and HCs, we conducted a study-wise genome-wide association study (GWAS) (Supplementary Fig. 16 and 17). Correlations between summary statistics from each study GWAS were estimated and studies with closer age alignment of participants showed higher genetic correlations (Fig. 4A), suggesting a gradual transition of genomic contributions to SUD over time. When compared with another published GWAS of SUD risk factors (Addiction risk factor; Addrf)^46,47^, genetic correlations between summary statistics in the corresponding GWAS were observed to increase with age and significant genetic correlation (*R^2^* = 0.19, *P* =7.16×10^-22^) was obtained between the UKB GWAS and published GWAS.

**Fig. 4.**
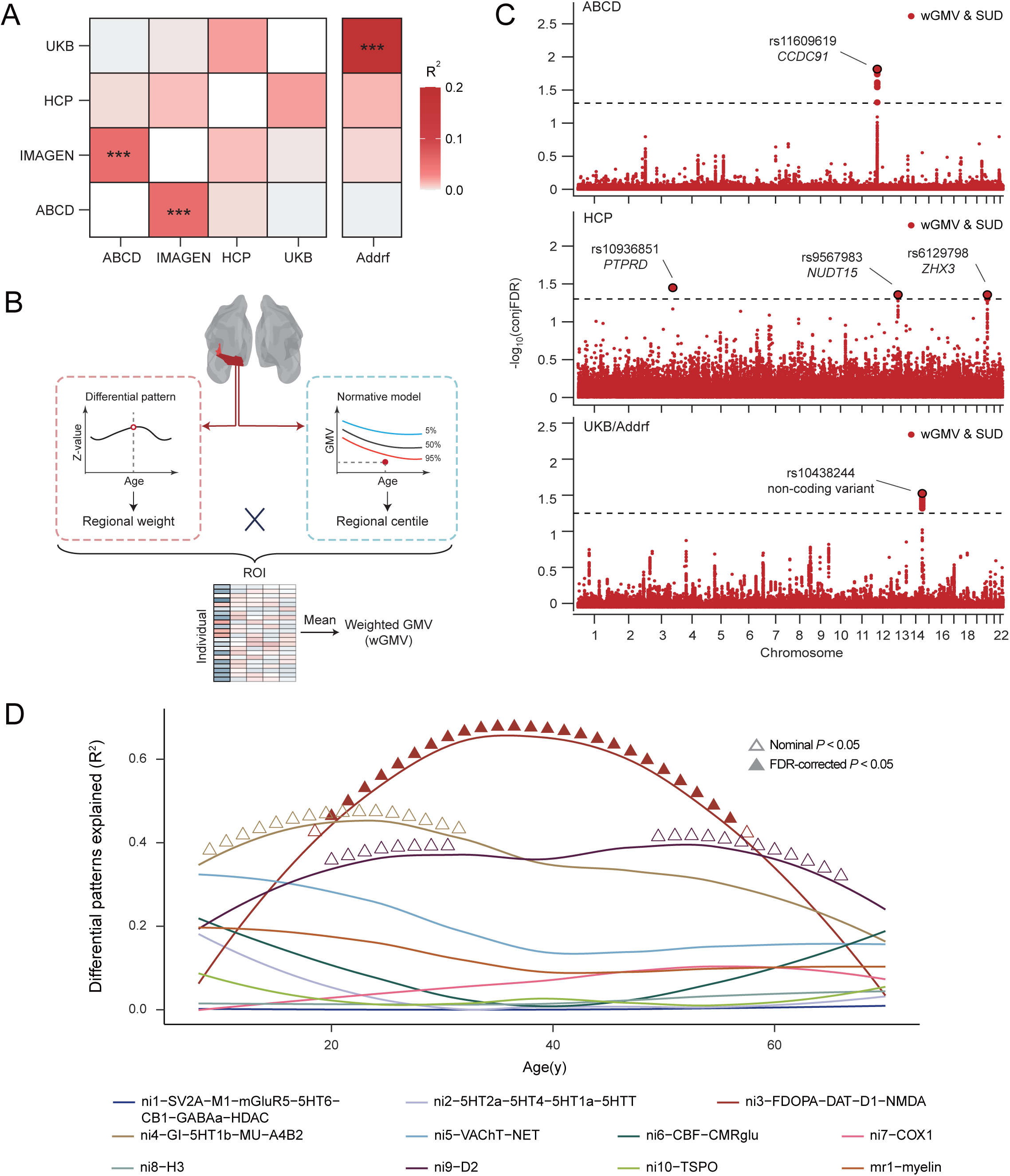
Comparison of genomic differences between HCs and SUDs across studies. (A) Genetic correlations between genetic variants associated SUDs in different studies; Addrf, addiction risk factor. *** <0.001. (B) Flowchart of weighted GMV (wGMV) calculation. wGMV was calculated by weighting thee region of interest (ROI) centiles with z-values from the corresponding SUD differential regional patterns and could be interpreted as a brain score reflecting the likelihood of SUD. (C) Conjuctional False Discovery Rate (FDR) manhattan plot for SUDs and SUD-related brain score in ABCD, HCP and UKB/Addrf. One significant locus rs11609619 on chromosome 12 (mapped to *CCDC91)* was identified in ABCD. Three significant loci rs10936851 on chromosome 3 (mapped to *PTPRD*), rs9567983 on chromosome 13 (mapped to *NUDT15*) and rs6129798 on chromosome 20 (mapped to *ZHX3*) were identified in HCP. Additionally, one significant locus rs10438244 on chromosome 14 was identified in UKB/Addrf. (D) Cortical volumetric differences between SUDs and HCs explained by neurobiological markers and cortical microstructure markers. Dimensionality-reduced molecular brain atlase were obtained from Lotter et al.. ▴: BH-FDR-corrected *P*<0.05; Δ: nominal *P*<0.05.

Next, a weighted-GMV (wGMV) score was calculated for each participant by weighing all ROI centiles with corresponding z-values obtained in the regional differential pattern for SUD (Fig. 4B), which represented one’s probability of being SUD (versus HC). Significant wGMV differences were observed between SUDs and HCs in all studies except HCP (ABCD: *d* = 0.05, *P* = 0.041; IMAGEN: *d* = 0.11, *P* = 0.026; HCP: *d* = 0.02, *P* = 0.196; UKB: *d* = 0.23, *P* = 1.64×10^-28^; one-sided t-test).

Finally, conjFDR was utilized to identify the localization of shared genetic variants between SUD and wGMV, localizing those specifically associated with SUD-related brain changes. At conjFDR < 0.05, five shared genetic loci were separately identified across the lifespan in ABCD, HCP and UKB/Addrf (Fig. 4C), whereas no significant locus was found in IMAGEN (Supplementary Fig. 18). During pre-adulthood (in ABCD), the shared locus rs11609619 on chromosome 12 was mapped to *CCDC91* and achieved significantly higher polygenetic score (PGS) for *CCDC91* expression in SUDs compared to HCs (*d* = -0.08, *P* = 0.007). *CCDC91* has been reported to be involved in the development and maintenance of white matter microstructure^48^, an essential component of brain tissue for efficient neural communications^49^ and neurodevelopment^50^. It was also found to be involved in several neurodevelopmental disorders in previous studies^51^, indicating an important role of brain maturation in the initiation of SUD. During early-to-mid adulthood (in HCP), three loci (rs10936851 on chromosome 3 mapping to *PTPRD*, rs9567983 on chromosome 13 mapping to *NUDT15,* and rs6129798 on chromosome 20 mapping to *ZHX3*) were identified, with only rs10936851 achieving significantly higher PGS for PTPRD (*d* = 0.15, *P* = 0.046) in SUDs. PTPRD is highly expressed in brain and works as a synaptic specifier and neuronal cell adhesion molecule^52^. Previous studies have implicated its involvement in dopaminergic reward pathway and increased vulnerability to substance addictiony^53,54^. Finally, during mid-to-late adulthood (in UKB/Addrf), the shared locus rs10438244 on chromosome 14 was identified, which is a non-coding transcript variant and was previously reported to be associated with alcoholism and tobacco use disorder^55^. Although it has been found to be associated with the expression of multiple genes^56^ (i.e. *TDRD9*, *KLC1* and *COA8*), only significantly higher PGS for *COA8* was observed in SUDs (*d* = -0.05, *P* = 0.011). *COA8* (previously referred to as *APOPT1*) plays critical role in the release of cytochrome *c* and protection against oxidative stress^57^, and has been indicated in the development of a broad spectrum of psychiatric disorders^58,59^. Combined with the lifespan trajectory for the effects of these loci on SUD (Supplementary Fig. 19), these findings suggest distinct genomic foundations for GMV-predicted SUD across different life stages.

To further examine the dynamic relationship between neurobiological markers and SUD-related volumetric differences, we performed a longitudinal association analysis leveraging the cortical gene expression atlas^60^ (Fig. 4D), which provided 20 molecular- and cellular-level markers alongside one cortical microstructure marker. During early-to-mid adulthood, dopaminergic neurotransmitter (ni3-FDOPA-DAT-D1-NMDA) significantly explained the estimated SUD-HCs brain differences in cortical ROIs, with the strongest effect observed around 34y. This result is consistent with the mechanistic pathway we identified within similar age range using HCP.

## Discussion

SUD has long been a critical public health issue due to its high prevalence and negative impacts on both mental and physical health^5,7,11,12^. Animal models and human studies have shown the importance of understanding the neurobiological mechanisms underlying the initiation and development of SUD^40,61^. However, due to the variability in brain morphology measurements and dynamic changes in volumetric GMV during different life stages, age has long been suggested as a potential confounder in elucidating the roles of neuroplasticity in SUD^23^. Here, by harmonizing neuroimaging, neurobehavioral and genomic data across multiple large population cohorts, we explored the lifespan trajectories of volumetric brain morphology associated with SUD, SUD-related neurobehavioral performances, and its genetic basis. The entire lifespan can be divided into three stages (Fig. 5): before 25y, where imbalanced regional brain development lead to SUD and SUD-related impulsivity; 25y to 45y, with enlarged ROIs in salience network and reinforcement of craving behaviors; after 45y, where SUD ultimately lead to widespread brain volumetric reductions.

**Fig. 5.**
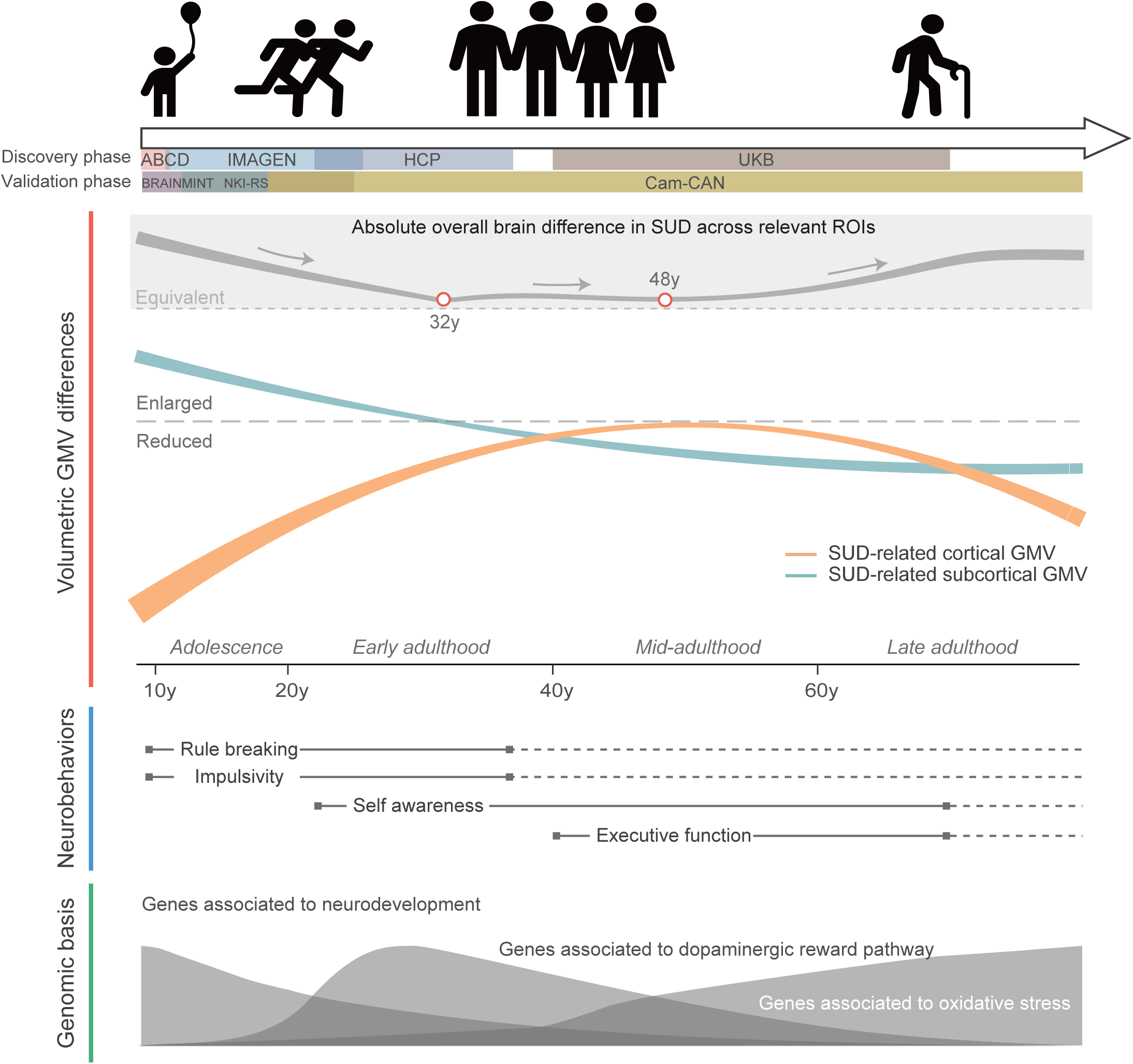
A panoramic view of dynamic changes in SUD encompassing brain structure, cognitive and behavioral performance, and genomic basis. The absolute overall brain difference in SUD across all ROIs was estimated by merging the p-values for cortical and subcortical differences and then converting the result into a Z statistic, while ignoring the directionality.

During pre-adulthood (from 8y to early 20s), individuals with SUD exhibited higher GMV in putamen, which may imply a greater propensity to habit learning in SUDs^41^, and lower GMV in nucleus accumbens, ACC, superior frontal and prefrontal cortex. Similar volumetric GMV differences in these brain regions were observed in participants with potential risk of developing follow-up SUD in the longitudinal analysis. Aligned with previous studies^26^, SUDs in IMAGEN only showed significantly decreased volumes in left medial orbitofrontal, which is more restricted compared to the brain regions in younger cohorts. This is likely due to the small sample size and SUD-associated neuroadaptations^62^ among IMAGEN adolescents. Our findings on brain regions with decreased SUD-related volumetric GMV (i.e. prefrontal, ACC and nucleus accumbens) were consistent with previous research on adolescent substance use^63–68^, suggesting important role of these ROIs on the development of adolescent SUD. However, although some studies have demonstrated evidence of larger dorsal striatum volumes for SUDs during early-adulthood^69–71^, few studies exist for adolescents. Our results extended existing findings on SUD initiation in pre-adulthood, and future research with larger sample sizes on adolescent SUD are required to provide further validation.

Combing the lifespan volumetric GMV and neurobehavioral patterns between SUDs and HCs, multiple potential mechanisms could be inferred to contribute to the development of SUD during adolescence. Firstly, developmental imbalance between dorsal striatum and prefrontal cortices was associated with impulsivity, which was also referred to as stopping impulsivity^72^, where dorsal striatum received extensive dopaminergic innervation from substantia nigra^73^ and were enlarged when the dopaminergic neurons are over-expressed in the nigrostriatal pathway^74^. It has been shown to be involved in a dopamine-related impulsivity trait^75^. This aligned with the findings from longitudinal analyses, indicating that impulsivity precedes substance use initiation, suggesting a potential causal role. Additionally, lower volumetric prefrontal cortices and ACC may be associated with lower top-down cognitive control, leading to increased rule-breaking behaviors during adolescence^76,77^. Consistent with previous findings^78,79^, our results demonstrated negative correlations between nucleus accumbens/ventral striatum and rule-breaking scores. This can be explained since nucleus accumbens receives projections from both the insula and the ventromedial prefrontal cortex^80^, and rule-breaking was associated with decreased activation of insula during reward anticipation^79^. Ventral striatum also plays an important role in adjusting goal-directed behavior based on predictions of future events, with action value signals broadcasted to prefrontal control regions to guide decision making^81^. This process is especially critical during adolescence, a period characterized by changing environmental demands and a growing need for peer connection, making adolescents particularly vulnerable to peer influence and subsequent substance use^82–84^. Taken together, these findings, along with other research on diverse cognitive differences in adolescent alcohol misusers^85^, underscore the complex and multifaceted nature of SUD development in this critical developmental period.

In addition, our results revealed negative correlations between the dorsal striatum and rule-breaking behaviors. As impairments in putamen and caudate function have been reported to be associated with inferior performances of rule-based tasks^86,87^, a possible explanation is that lower volumes of putamen and caudate lead to dysregulation of rule-based behavior. Our findings on the genomic basis of SUD-related neurobehavioral changes also highlighted the importance of neurodevelopment, with gene *CCDC91* identified to be involved in the onset of neurodevelopmental disorders.

Significantly, brain volumetric differences between SUDs and HCs in pre-adulthood diminished when transitioning from late adolescence to early adulthood. Given that adolescence is a period characterized by morphological and functional transformations of the brain^88^, we hypothesize that the diminishing differences between SUDs and HCs could be explained by continued neurodevelopment or by compensatory adjustment of brain function. Longitudinal investigation suggested a potential mechanism of imbalanced neurodevelopment in ROIs between subcortex and cortex for SUD. Therefore, the resilience-like volumetric changes during early life stage could be explained either by the immature brain development in the absence of substance use^89^, or neuroadaptations of the reward system in the presence of substance use^20^. Studies using rodent models demonstrated that enriched environment during adolescence could reduce drug sensitivity in adulthood by decreasing the dopaminergic neurons in substantia nigra and activity of the hypothalamic-pituitary-adrenal axis^90–94^. This suggested a unique adaptation of the brain to dopamine-induced behaviors in adolescence and a possible crucial interventional window for SUDs before the age of 30.

Increased volumetric GMV was observed in SN for participants with SUD in early-to-mid adulthood, specifically in rostral ACC, insula and lateral orbitofrontal. This network is known to be involved in emotion, self-awareness, autonomics^95^, and plays a role in mediating conscious emotion and interoception associated with addictive substances^96^. The increased regional GMV of SN might be attributed to hyperconnectivity within SN^97,98^ and enhanced craving behaviors^99,100^ associated with increased dopaminergic activity^101^. However, some studies showed inconsistent results with decreased GMV of insula and activation of multiple addiction associated tasks^102^, where inconsistency depend on the severity and duration of SUD. Due to the negative effect of overactivation of SN on mood sensitivities^103^, lower self-awareness and mood instability were observed. These factors are believed to contribute to the substance craving behaviors and self-administriation^101,104^. Compared to the complex cognitive differences observed in early adulthood SUD, only that factor encompassing general cognitive impairment, including inhibitory control, and memory processes was linked to SUD in mid-to-late adulthood, as revealed by brain-informed correlation analysis. Remarkably, these findings are the first to lend support from the perspective of population analysis to the hypothesis that, as drug experience accumulates, the characteristics of drug use shifts from impulsive behaviors to compulsive behaviors^41,44^. Alongside this, the more general cognitive impairment in mid-to-late adulthood might be explained by the progressive expansion of drug-related brain activity from the ventromedial prefrontal regions toward more posterior and even temporal regions^105^, suggesting an important additional burden on the management of elderly drug users due to general intellectual deterioration which will require additional rehabilitation.

Further, identification of a significant SUD-associated *PTPRD* locus suggested potential overactivation of the dopaminergic reward pathway and enhanced values of additive substance cues. This result was also supported by an age-related increase in the explained variance of a dopaminergic factor to the brain change within the age range for HCP participants. Specifically, this factor was mainly composed of dopamine precursors FDOPA, transporters DAT, D1 receptors enhancing drug-seeking behavior, and NMDA receptors involved in habit formation. Therefore, early-to-mid adulthood can be characterized by addiction development with positive feedback related to awareness/mood and reward system. It should be noted that the volumetric differences between SUDs and HCs could only be observed for alcohol and marijuana users, and future research of larger sample is called for smoking and intra-venous drug administration^106^.

The SUD-associated volumetric changes of SN diminished during mid-to-late adulthood, which could be explained by decreased functional connectivity and activation among older adults^107,108^, increased reward threshold^62^, and potential neurotoxicity of the substances. This was subsequently supported by the findings from the genetic analysis that significant SUD-associated locus *COA8* was involved in the oxidative stress process. Brain regions with significant volumetric differences between SUDs and HCs emerge from subcortex, amygdala, hippocampus, nucleus accumbens, extend to superior frontal and frontal pole cortex, and finally to orbitofrontal cortex, naturally aligning with the timing of brain development and highlighting the importance of neurodevelopment during the onset of SUD. Finally, the age-specific genetic correlations across different studies additionally underscore the gradual evolving process of SUD-related mechanisms from a genetic perspective.

Regarding hemispheric differences, although similar lifespan changes in both hemispheres were observed, regions related to top-down response inhibitory control appeared to be more pronounced in the right hemisphere.

While the homogeneity in the brain differential patterns across different substances was found, this was not the case for participants with drug use disorder in the subgroup analysis, particularly in terms of late adulthood. Despite the evidence that some types of drugs may cause gliosis, leading to larger regional volumes^109,110^, this result might arise from selection bias due to our exclusion criteria for participants with neuropsychiatric disorders and a very small relative sample size in the drug use disorder in UKB.

To the best of our knowledge, this is the first study exploring the dynamics of SUD associated volumetric trajectories across the lifespan. Our study demonstrated the importance of brain maturation in elucidating the neurobiological mechanisms and genetic basis contributing to SUD. Nonetheless, there are several limitations associated with this study. Firstly, due to limited accessibility of DSM assessment, definition of SUD mostly depended on self-reported frequency and quantity of substance use. It should come to mind that since ABCD does not include questions related to the substance use experience, the definition of SUD has had to be relaxed to include substance misuse behaviors—that is, any over-early use of addictive substances for participants. While this may seem to be a relatively broad definition, only 0.7% of individuals in ABCD are identified as SUDs under this criterion. Secondly, although validated using longitudinal data, the lifespan GMV trajectories for SUDs and HCs were estimated using cross-sectional data, with limited sample sizes in childhood and mid-to-late adulthood. Further validations are needed for early adulthood, where a noticeable shift of brain mechanisms could potentially occur. Cohorts with large sample sizes and more detailed substance use data as well as actual clinical diagnosis are needed to validate the robustness of our findings. Thirdly, different neurobehavioral measurements were adopted in different studies, which could lead to bias in identifying brain regions associated with SUD. Although we tried to tackle this issue by extracting common factors for rule-breaking behaviors and impulsivity across studies, consistent neurobehavioral measurements are called for more accurate identification of consistent SUD-associated brain regions. Fourthly, while previous studies have offered rationales for using cross-sectional brain atlases in longitudinal association analysis^60^, bias is inevitable, particularly for age-sensitive research. Finally, caution should be taken when interpretating consistently higher impulsivity among SUD participants, mostly in self-reported measures. Given that impulsivity is considered a broad and multifaceted concept, different test results may convey different messages. Future research on multiancestry comparisons is also called for more generalized conclusions.

## Methods

### Ethical statement

All cohort data used in this study comply with relevant ethical regulations and informed consent was sought from all participants and a parent/guardian of each participant if under 18 years in all studies. ABCD and HCP study was supported by the National Institutes of Health (NIH). The IMAGEN study was approved by local ethnical research committees at each research site: King’s College London, University of Nottingham, Trinity College Dublin, University of Heidelberg, Technische Universitat Dresden, Commissariat a l’Energie Atomique et aux Energies Alternatives, and University Medical Center. UK Biobank has approval from the North West Multi-centre Research Ethics Committee as a Research Tissue Bank approval. NKI-RS received approvals from the Nathan Kline Institute and Montclair State University. Cam-CAN was approved by the local ethics committee, Cambridgeshire 2 Research Ethics Committee and BRAINMINT was approved by the Regional Committee for Medical and Health Research Ethics South-Eastern Norway.

### Participants

Participants from four large population cohorts (ABCD, IMAGEN, HCP and UKB) were harmonized to estimate the lifespan trajectories of volumetric GMV for SUDs and HCs in the discovery set, and those from NKI-RS and Cam-CAN were harmonized similarly for external validation except BRAINMINT. In all studies, participants with both non-missing neuroimaging and substance use data were included, and those with any serious medical or neurological conditions, pre-existing psychiatric or neuropsychiatric disorders other than SUD, other illnesses that could confound neuroimaging, or with GMV beyond 4 interquartile ranges in any ROI were excluded.

ABCD is a longitudinal neuroimaging cohort with 11,875 adolescents from 21 sites across the United States enrolling at baseline. The version 3.0 (baseline; 9-10 years old) and 4.0 (follow-up; 11-13 years old) of the annual curated data releases for ABCD (https://abcdstudy.org/about/) were used in this analysis. At baseline, a total of 11,301 participants (52.2% males) consisting of 11,217 HCs and 84 SUDs were included, with 7,775 having follow-up imaging visits. IMAGEN is a multicenter neuroimaging longitudinal cohort, where approximately 2,000 healthy adolescents of European descent across multiple sites in Europe were recruited at baseline (age 14y) and multiple follow-up visits at 16y (FU1), 19y (FU2) and 23y (FU3). A total of 1,504 participants (53.7% males) with 1,298 HCs and 185 SUDs at baseline, 539 HCs and 419 SUDs at FU2, and 345 HCs and 450 SUDs at FU3 were included in this analysis, with average number of MRI scans per participant being 2.15. HCP is a cross-sectional cohort of healthy young adult aged 22y – 37y and a total of 1,113 participants (45.6% males) with 396 HCs and 310 SUDs were included in this analysis. UKB is an ongoing population cohort with over 500,000 participants aged 37y – 73y recruited in 2006-2010, where follow-up neuroimaging assessments were applied to a subset of participants from 2014 to 2019. A total of 37,549 participants (47.3% males), with 6,243 HCs and 5,897 SUDs at baseline and 914 HCs and 328 SUDs at follow-up visits were included in the analysis. NKI-RS is an ongoing cross-sectional cohort of adults in Rockland County^111^, and a total of 1,112 participants (37.6% males) aged 12y – 80y with 862 HCs and 250 SUDs were included for external validation. Cam-CAN is a cross-sectional project designed to explore healthy aging, with 700 participants in UK aged 12y – 89y examined for brain structure measures^112^, and a total of 445 participants (49.0% males) with 140 HCs and 305 SUDs were included for validation. The BRAINMINT study is a longitudinal study investigating the mechanisms of brain plasticity involving children and adolescents aged 9y – 25y from Oslo in Norway. Only cross-sectional data from 570 participants (28.4% males) were included for the validation analysis, comprising 414 HCs and 156 SUDs. Detailed descriptions of population characteristics for studies were provided in the Supplementary Tables 1 and 9.

### Assessment of substance use

Due to the varying accessibility of addictive substances for participants in different life stages, different standards were used to identify participants with SUD and HC. For ABCD, according to the self-reported substance use interview records, SUD was defined as participants with: 1) any full drink of beer, wine or liquor; 2) more than just a puff of cigarette, electronic cigarette, “chew”, cigar, hookah, pipes, and nicotine replacement; 3) more than just a puff of marijuana-included products and fake marijuana; and 4) any try of stimulants, depressants, hallucinogens and inhalants, and HC was defined as those with no use of any addictive substance. For IMAGEN, SUD was defined as participants with: 1) child Alcohol Use Disorders Identification Test (AUDIT) total score > 7; 2) Fagerstrom Test for Nicotine Dependence (FTND) score >= 1; 3) any try of marijuana (grass, pot) or hashish for participants at baseline / > 10 times for participants at follow-ups; and 4) any try of stimulants, depressants hallucinogens and inhalants, and HC was defined as those with no exposure to any addictive substance. The frequency and quantity of substance use was measured by European School Survey Project on Alcohol and Drugs (ESPAD) questionnaire. For HCP, SUD was defined as participants with: 1) DSM-4 criteria for alcohol abuse or alcohol dependence met; 2) FTND score ≥ 4; 3) DSM-4 criteria for marijuana dependence met; and 4) any use of illicit drugs > 10 times, and HC was defined as those meeting all of the following requirements: 1) no DSM-4 alcohol dependence or abuse symptoms endorsed; 2) no smoking history or only experimental uses; 3) no marijuana used ever; 4) no use of illicit drugs ever. For UKB, SUD was defined as participants with: 1) weekly alcohol assumption > 15 units and alcohol use almost daily; 2) current smokers on most or all days; 3) cannabis ever taken > 100 times; and 4) ever addicted to illicit or recreational drugs, and HC was defined as those meeting all of the following requirements: 1) less than weekly use of alcohol; 2) no current and past smoking; 3) no marijuana use; 4) never addicted to illicit or recreational drugs. For NKI-RS, SUD was defined as participants with: 1) any use of tobacco, alcohol, marijuana and additive drugs (age<18); 2) clinical diagnosis of substance abuse or dependency in DSM-IV (age≥18), and HC was defined as those not meeting the criteria of SUD. For Cam-CAN, SUD was defined as participants with: 1) weekly alcohol use ≥ 3 or 4 times; 2) current smoking; 3) intermediate or substantial drug use severity, and HC was defined as those meeting all of the following requirements: 1) monthly alcohol use < 4 times or less or past drinker; 2) total smoking quantity < 100 or past smoker; 3) no drug used ever. For BRAINMINT, substance use behaviors were measured using CRAFFT+N questionnaire. SUD was defined as participants with any use of full drink containing alcohol, tobacco-related products, marijuana or anything else to get high in past year and CRAFFT score >= 2, and HC was defined as those not meeting the criteria of SUD.

In sensitivity analysis, we tested whether the discontinuity of brain volumetric differences between SUDs and HCs from pre-adulthood to adulthood could be attributed to the change of SUD/HC definition. Definitions of SUD and HC were modified by replacing all above defined HCs in HCP and UKB as mild SUDs.

### Assessment of behavioral and neurocognitive performances

Neurobehavioral measurements were selected according to potential circuits associated with SUD^39,40^. In ABCD, rule breaking and conduct problem scores from Child Behavior Check List (CBCL), “fun seeking” score from Behavioral inhibition and Behavioral Activations scales (BIS/BAS), total impulsivity score from the UPPS-P Impulsive Behavior Scale, Flanker Inhibitory Control and Attention Test (Flanker) and Stop Signal Test (SST) were used for the measurement of rule-breaking behaviors and impulsivity. The accuracy rates of small reward trials and large reward trials adjusted the accuracy in neural trials from Monetary Incentive Delay task (MID) were used to measure reward sensitivity. The Dimensional Change Card Sort Test (DCCS) score was used to measure the cognitive flexibility aspect of executive function. In IMAGEN, conduct problem score from Strengths and Difficulties Questionnaire (SDQ), novelty seeking personality score from Temperament and Character Inventory (TCI), average sensation seeking and impulsivity scores from Substance Use Risk Profile Scale (SURPS) as well as Monetary-Choice Questionnaire (KIRBY) were used for the measurement of rule-breaking behaviors and impulsivity. The CANTAB Spatial Working Memory (SWM) test was used to measure working memory components of executive function^113^. In HCP, rule breaking score from Achenbach Adult Self Report (ASR), conduct problem score from Semi-Structured Assessment for the Genetics of Alcoholism (SSAGA), Flanker test as well as delayed discounting task were used to measure rule-breaking behaviors and impulsivity. The index in discounting task is the Area Under the Curve for Discounting of different delayed reward vary in $200 and $4K. A smaller AUC indicates steep delay discounting, i.e., increased preference toward short-term benefits. DCCS was used for the measurement of executive function. We also included emotional measurements offered by NIH Toolbox, where participants were asked to evaluate their life experiences in terms of life satisfaction, meaning and purpose, and perceived stress. In UKB, the only question about rule-breaking is whether people treated themselves as a risk-taking person. Other measurements for psychological well-being included mood swing, irritability, fed-up feelings, anhedonia feeling and life satisfaction for individuals. Executive function was measured by numeric short-term memory task and tower rearranging test. In NKI-RS, the health subscale risk taking score from the Domain-Specific Risk-Taking Scale (DOSPERT), rule-breaking behavior score from youth/adult self-report (YSR/ASR), sensation seeking score from UPPS-P Impulsive Behavior Scale, impulsivity score from parent-rated Conners ADHD Rating Scale (Conner-P) and Conners Adult ADHD Rating Scale (CAARS), social awareness score from Social Responsiveness Scale (SRS), executive functioning score from Conner-P and conflict effect score from Attention Network Test (ANT) were used. Self-esteem score was calculated as the sum of the 7^th^, 8^th^ and 14^th^ term in Beck Depression Inventory (BDI) questionnaire.

### Assessment of adverse childhood experience

The total childhood trauma experiences score was calculated as the number of reported events at initial measurements across emotional neglect, physical neglect, emotional abuse, physical abuse and sexual abuse subscales. In ABCD, emotion neglect from Children’s Report of Parental Behavioral Inventory (focused on the first caregiver; ≥2 matching descriptions), physical neglect from Parent Monitoring Survey (focused on the first three personal safety questions; ≥1 matching descriptions), abuse (764-765 for emotional abuse; 761-763 for physical abuse; 767-769 for sexual abuse) from Parent Diagnostic Interview for DSM-5 (KSADS) Traumatic Events - post-traumatic stress disorder module (≥1 matching descriptions) were obtained as suggested^114^. In IMAGEN, all subscales were obtained from Childhood Trauma Questionnaire (CTQ) with pre-defined cut-off values^115^.

### Acquisition, imputation and quality control of genomic data

For ABCD, the genotype data were obtained directly from the public release 3.0, and imputed using the Michigan Imputation Server with hrc.r1.1.2016 reference panel^116^ and Eagle v2.3 phasing. Due to genetic diversity85 and low linkage disequilibrium (LD) levels of African populations86, a total of 2,387 ABCD subjects self-reporting ancestral origins as Black or African American was excluded. One participant per family was randomly selected to avoid within family correlations. For IMAGEN, details of the genotyping and quality control could be found in Desrivières et al^117^, imputed using the TOPMed imputation server with the HapMap3 reference panel^118^. For HCP, detailed genotyping and quality control could be found in^119^, where imputation was performed with the 1000 Genomes panel^120^. For UKB, detailed genotyping and quality control procedures are available in^121^. Individuals that were estimated to have recent British ancestry and have no more than ten putative third-degree relatives in the kinship table were included. Similar quality control standards were performed by PLINK 1.90 across studies. Individuals with >10% missing rate and single-nucleotide polymorphisms (SNPs) with call rates < 95%, minor allele frequency < 1%, deviation from the Hardy-Weinberg equilibrium with P < 1E-10 were excluded from the analysis. Thus, a total of 7,662 participants and 5,020,358 SNPs in ABCD, 1,982 participants and 5,966,316 SNPs in IMAGEN, 897 participants and 5,375003 SNPs in HCP, and 337,151 participants and 8,894,431 SNPs in UKB were included in the genomic analysis.

### Acquisition and preprocessing of neuroimaging data

Neuroimaging data in ABCD were obtained using 3T scanners (Siemens Prisma, General Electric MR750 and Philips Achieva dStream) with 32-channel head coil and high resolution T1-weighted structural MRI. The pre-processing processes and quality control procedures were completed by the ABCD research teams according to the ABCD standard pipeline and protocol^122,123^. Neuroimaging data in IMAGEN were obtained using 3T MRI systems based on the ADNI protocol from 4 different manufacturers (Siemens Philips, GE Healthcare, and Bruker), with detailed MR protocols and QC procedures described in^124^. In HCP, neuroimaging data were obtained on a Siemens Skyra 3T scanner employing a 32-channel head coil with protocols provided in https://www.humanconnectome.org. In UKB, neuroimaging data were obtained with a standard Siemens Skyra 3T scanner with a 32-channel head coil, identical in both hardware and software in Manchester, Newcastle, and Reading. Details of preprocessing and QC processes could be found in^125^. In brief, quality-controlled processed T1-weighted neuroimaging data were obtained directly from cohort teams. The NKI-RS neuroimaging data were obtained following the same protocols as HCP provided by the University of Minnesota. Quality controlled T1 images were downloaded. The Cam-CAN neuroimaging data were provided by the University of Cambridge using the same MRC CBU scanner according to^126^. The BRAINMINT neuroimaging data were obtained using a 3T GE SIGNA Premier scanner with a 48-channel head coil. Then regional volumes were extracted by FreeSurfer v6.0 (BRAINMINT processed by v7.3.2) cross-sectional pipelines using Desikan-Killiany (h.aparc) atlas for cortical regions, and ASEG atlas for subcortical regions, except for UKB, where Brain volumetric phenotypes could be acquired directly by category ID 192&190. Quality check was performed according to FreeSurfer reconstruction quality-controlled (QC) measures.

### Harmonization of neuroimaging data across multiple cohorts via GAMLSS

Motivated by the use of GAMLSS model in construction of brain charts^32^, normative model was used to harmonize neuroimaging data across multiple study cohorts. ComBAT method was first employed to remove confounding due to imaging scanners within each study (*ComBat* function in sva-3.46.6 package). For Cam-CAN, due to a change in the scanner coil during data collection and differences in MTI TR across participants, these two confounding variables were also preemptively corrected using ComBAT. Next, regional GMV was modeled using a generalized gamma distribution, with mean and variance defining functions involving fractional polynomials of age, sex, handedness and study-specific random effects (gamlss-5.4.18 package)^32^. The number of fractional polynomials and whether to include a study random effect was determined for each ROI based on Bayesian information criterion (BIC) (Supplementary Fig. 2). The same optimal GAMLSS models were also applied during the validation phase. Only cross-sectional HCs were used for model fitting, including baseline ABCD data, sampled IMAGEN data, HCP data and baseline UKB data. A single observation was randomly sampled from all visits among IMAGEN Individuals with more than one neuroimaging scans, with sampling probability being standardized reciprocal of the distribution frequency. Finally, volumetric characteristics for each participant were placed on the normative model to obtain age- and sex-specific centiles.

### Estimation of the lifespan volumetric GMV trajectories for SUDs and HCs

Since GAMLSS does not provide standard error estimates, generative additive model (GAM) was used to compare mean regional GMV across different ages between SUDs and HCs. First, confounding effects due to sex, handedness and study on the GMV measures were removed, and covariate-adjusted GMV were obtained, with the underlying assumption that effects of these confounders on GMV were independent of substance use status. Next, lifespan GMV trajectories across age were constructed separately for SUDs and HCs using GAM (*gam* function in mgcv-1.9.0). Cubic spline with *K*=3 was used as the basis function for age (spline-4.2.2). To obtain continuous comparisons of the age-dependent GMV trajectories between SUDs and HCs, we began by predicting the mean and standard error estimated from group-level GMV trajectories at discrete ages between 8y and 70y with a step of 0.05y. Two-sided z test was used to test the null hypothesis that population GMV in SUDs and HCs was identical. After correcting for multiple comparisons via False Discovery Rate (FDR) method, GAM with cubic spline basis were selected to construct continuous significance comparisons between SUDs and HCs across the lifespan. Then, we explored whether there was heterogeneity in brain differential patterns for SUDs from two aspects. On one hand, SUDs were divided into four groups with different substance use behaviors, including alcohol, tobacco, marijuana and any other type of addictive drugs. Lifespan volumetric patterns were estimated separately for these four groups and compared with HCs. On the other hand, hierarchical clustering method was used for individual GMV centiles to investigate potential clustering characteristics in changing patterns between brain regions for SUD.

### Validation of the lifespan volumetric GMV trajectories for SUDs and HCs

The lifespan brain differential trajectories for SUD were validated in two ways. Firstly, validation was performed using the longitudinal ABCD and UKB datasets, where the baseline data overlapped with the training data. Individual longitudinal volumetric changes were calculated as the difference in GMV between follow-up and baseline. The change rate in SUD relative to HC was obtained using the linear regression model adjusting for age and sex. The magnitude and direction of change rates for ROIs were compared with the estimated slope differences at the corresponding individual ages using fitted GAM models. following the same estimation process and parameter settings as mentioned above. Second, external NKI-RS and Cam-CAN datasets were harmonized following the same estimation process and parameter settings as mentioned in the discovery analysis above, resulting in an independent normative model for the entire validation phase. Since the BRAINMINT samples could only be analyzed on a local server, an inverse-variance weighted meta-analysis was conducted to combine the HC and SUD change trajectories from BRAINMINT with those derived from NKI-RS and Cam-CAN samples in overlapping ages. Then two-sided z tests at different time points and GAM with cubic spline basis were used to refit the differential trajectories similarly. As it is hard to obtain statistically significant results in the validation sets due to limited samples, spearman correlation analysis was used to assess the consistency of differential patterns in the discovery and validation datasets.

### Comparison of lifespan neurobehavioral performances between SUDs and HCs

Two-sided t-test was used to compare neurobehavioral measurements between SUDs and HCs in the discovery phase. In ABCD, we also applied down-sampling methods to address the severe class imbalance problem in SUD. A linear interaction term between age and SUD was included to evaluate the time-varying neurobehavioral changes in SUDs in external validation, with age, SUD, sex, handedness included as covariates. Standard error was calculated assuming multivariate normal distribution. Factor analysis was applied to calculate composite scores for rule-breaking and impulsivity due to heterogeneity of measurement tools (Supplementary Fig. 13). As there is only one self-reported question about risk-taking personal characteristics related to rule-breaking or impulsivity in UKB, it is not included in the brain-neurobehavior correlational analysis. The number of common factors were determined by the number of eigen values larger than 1. Correlations between the extracted factors and GMV centiles of each ROI were calculated for all studies. BH-FDR method was used for multiple testing. To highlight the impact of lifespan brain-neurobehavior correlations, effect sizes were meta-analyzed within studies of adolescents (ABCD and IMAGEN), whereas HCP was only used as the resource of adult study. Since rule-breaking and impulsivity are mildly correlated in ABCD (*r* = 0.08, *P_adj_* = 1.47×10^-12^), we adjusted for rule-breaking score in the correlational analysis between impulsivity and brain GMV, and vice versa as a sensitivity analysis.

### Longitudinal association analysis using brain atlases

Cognitive term based meta-analysis statistical brain atlases^43^ were downloaded from Neurosynth^43^ (https://neurosynth.org/) and parcellated according to the APARC and ASEG atlases using NiMARE 0.0.11^127,128^. Factor analysis with varimax rotation was then used. All factors explaining > 1% of the variance were kept, resulting in seven factors. To ensure that these factors were highly representative of underlying cognitive processes, only cognitive terms with loadings > 0.2 and also within the top 50^th^ percentiles were retained for each factor. Factor loadings and factor-level brain atlases are available in the Supplementary Fig. 11-12. Subsequently, univariate correlation analyses were performed between the factor-level cognitive brain atlases and differential z-statistic of ROIs between SUD and HC at 0.5y intervals from 8y to 70y. Explained variance (*R^2^*) was calculated to represent the influential levels, and *P*-values were determined using 1000 spin permutations. BH-FDR method was used for multiple testing within each ROI. A similar analysis was conducted using neurobiomarker brain atlases^60^, where 20 molecular- and cellular-level factors were obtained along with one cortical microstructure factor from cortical gene expression atlases, limiting the analysis to cortical region of interest. *P*-values were also calculated via 1000 permutations and adjusted for multiple testing by BH-FDR method, with null maps generated according to^60^.

### GWAS, genetic correlation analysis and conjFDR

Scalable and accurate Implementation of generalized mixed model^55^ (SAIGE) was used for genetic associations to avoid inflating Type I error due to case-control imbalance, and *P* values were obtained via saddlepoint approximation. Given the longitudinal nature of ABCD and IMAGEN, participants were defined as cases when they can be classified as SUDs for more than the average times of the total visits. This resulted in 6,177 cases (SUDs) and 1,485 controls (HCs) in ABCD, and 605 cases and 1377 controls in IMAGEN, respectively. To better understand how estimated GMV trajectories may contribute to the development of SUD, we calculated a wGMV score, representing the possibility of being classified as SUD using GMV. This score was composed of all ROI centiles weighted by the z-value obtained in the GWAS comparing SUDs and HCs. When wGMV was used as the continuous phenotype, GWAS was conducted using Plink 2.0, with sex, handedness, age at baseline, self-reported ethnicity, and the top *k* population genetic principal components (*k* = 20 for ABCD and *k* = 10 for the other three studies) included as covariates. Within-sibship GWAS^129^ was used for both binary and continuous phenotype GWAS for HCP. A published GWAS of SUD risk factors^46,47^ (Addrf) was used as a reference to assess the consistency of our GWAS results, where cross-trait polygenic risk score was calculated and used to evaluate genetic correlations between summary statistics of the published GWAS and SUD GWAS. Specifically, the published GWAS was used as the testing sample and PGS was calculated on the testing sample after LD-based pruning (window size=50kb, variant count=5 and *R^2^* threshold=0.1) using the GWAS summary statistics from the current GWAS in Plink 1.90, adjusting for sex, handedness, age, self-reported ethnicity, and population genetic principal components. Estimated genetic correlations were then corrected using *bcPRS* function in R^130^.

Finally, ConjFDR^131^ was used to identify specific shared genetic variants between SUD and wGMV. Test statistic for the SNP effects on SUD is re-ranked based on their associational strength with wGMV, obtaining the conditional FDR value. The maximal conditional FDR value was selected as the conjFDR value. We also investigated common genetic variants shared between wGMV in UKB and the SUD risk factor with the study population primarily consisting of individuals in mid-to-late adulthood. As no significant results were found between SUD and wGMV in UKB (Supplementary Fig. 18), we instead reported the conjFDR results between SUD in UKB and the SUD risk factor.

Genetic variants identified in the above analysis were mapped to genes using Open Targets Genetics (www.opentargets.org), which integrates evidence from molecular phenotype quantitative trait loci, chromatin interaction, in silico functional predictions and distance between the variant and the canonical transcript start site of genes. To further prove the effects of specific genes on SUD, we calculated PGS for both RNA expression and protein level of corresponding genes based on publicly available GWAS summary statistics^132^. Two-sided t-test was used to compare the PGS between SUDs and HCs.

## Data Availability

The raw ABCD, IMAGEN, HCP, UKB, NKI-RS, Cam-CAN and BRAINMINT data are protected and are not available due to data usage agreement. However, access can be obtained upon application except BRAINMINT samples, which could only be analyzed on local server in the University of Oslo. ABCD data can be accessed at https://abcdstudy.org/; IMAGEN data can be accessed by email at https://imagen-project.org/; HCP data are available from: https://www.humanconnectome.org/; UKB data can be accessed at https://biobank.ndph.ox.ac.uk/; NKI-RS data can be applied by mail; and Cam-CAN data can be accessed from: http://www.mrc-cbu.cam.ac.uk/datasets/camcan/. Cognitive term and factor-level gene expression brain atlases can be accessed at https://neurosynth.org/ and https://github.com/LeonDLotter/CTdev/. Published GWAS summary statistics of SUD risk factor (Addrf) can be accessed by mail from^46^. Publicly available GWAS summary statistics for multi-omics traits can be accessed at https://www.omicspred.org/.

## Code Availability

Primary analyses were conducted in R v4.2.2 and atlas-related analysis were conducted in Python 3.9.10. GAMLSS models were performed using gamlss-5.4.18 and gamlss.dist 6.1-1 R packages. GAM models were built using mgcv 1.9.0 R package. NiMARE 0.0.11 was used to download and process Neurosynth atlases. SAIGE was used to perform GWAS (https://github.com/saigegit/SAIGE) and genetic correlation was calculated following codes from https://github.com/xm1701/bcPRS?tab=readme-ov-file using Plink 1.90 and bcPRS 0.0.0.9000 R package. ConjFDR was used to identify genetic loci shared between two phenotypes (https://github.com/precimed/pleiofdr). Open Targets Genetics was used to perform functional annotations (www.opentargets.org).

## Acknowledgments

We thank Professor Alexander S. Hatoum for providing the GWAS summary statistics of SUD risk factor. This work received support from the following sources: the General Projects of Shanghai Science and Technology Commission (21ZR1405000 [X.L.]), the National Nature Science Foundation of China (No.82304241 [X.L.]), National Key R&D Program of China (No.2018YFC1312904 [J.F.], No.2019YFA0709502 [J.F.] and No.2023YFE0199700[X.C.]), Shanghai Municipal Science and Technology Major Project (No.2018SHZDZX01 [J.F.], ZJ Lab [J.F.], and Shanghai Center for Brain Science and Brain-Inspired Technology [J.F.]), the 111 Project (No.B18015 [J.F.]), the China National Postdoctoral Program for Innovative Talents (BX20240086 [S.X.]), the Shanghai Yangfan Project (24YF2702200[S.X.]), the European Union-funded FP6 Integrated Project IMAGEN (Reinforcement-related behaviour in normal brain function and psychopathology) (LSHM-CT-2007-037286 [G.S.]), the UK Research and Innovation (UKRI) funded UK government’s Horizon Europe funding guarantee (10041392 and 10038599 [G.S.]), the Horizon 2020 funded ERC Advanced Grant ‘STRATIFY’ ((Brain network based stratification of reinforcement-related disorders) (695313 [G.S.]), Human Brain Project (HBP SGA 2, 785907, and HBP SGA 3, 945539 [G.S.]), the Medical Research Council Grant ’c-VEDA’ (Consortium on Vulnerability Externalizing Disorders and Addictions) (MR/N000390/1 [G.S.]), the National Institute of Health (NIH) (R01DA049238 [G.S.], A decentralized macro and micro gene-by-environment interaction analysis of substance use behavior and its brain biomarkers), the National Institute for Health Research (NIHR) Biomedical Research Centre at South London and Maudsley NHS Foundation Trust and King’s College London, the Bundesministeriumfür Bildung und Forschung (BMBF grants 01GS08152; 01EV0711 [G.S.]; Forschungsnetz AERIAL 01EE1406A, 01EE1406B; Forschungsnetz IMAC-Mind 01GL1745B [G.S.]), the Deutsche Forschungsgemeinschaft (DFG grants SM 80/7-2, SFB 940, TRR 265, NE 1383/14-1 [G.S.]), the Medical Research Foundation and Medical Research Council (grants MR/R00465X/1 and MR/S020306/1 [S.D.]), the National Institutes of Health (NIH) funded ENIGMA (grants 5U54EB020403-05 and 1R56AG058854-01 [S.D.]), NSFC grant 82150710554 and environMENTAL grant 101057429. Further support was provided by grants from: - the ANR (ANR-12-SAMA-0004, AAPG2019 - GeBra [J.-L.M.]), the Eranet Neuron (AF12-NEUR0008-01 - WM2NA; and ANR-18-NEUR00002-01 - ADORe [J.-L.M.]), the Fondation de France (00081242 [J.-L.M.]), the Fondation pour la Recherche Médicale (DPA20140629802 [J.-L.M.]), the Mission Interministérielle de Lutte-contre-les-Drogues-et-les-Conduites-Addictives (MILDECA [J.-L.M.]), the Assistance-Publique-Hôpitaux-de-Paris and INSERM (interface grant [M.- L.P.M.]), Paris Sud University IDEX 2012 [J.-L.M.], the Fondation de l’Avenir (grant AP-RM-17-013 [M.- L.P.M.]), the Fédération pour la Recherche sur le Cerveau; the National Institutes of Health, Science Foundation Ireland (16/ERCD/3797 [R.W.]) and by NIH Consortium grant U54 EB020403 [S.D.], supported by a cross-NIH alliance that funds Big Data Knowledge Centres of Excellence. ImagenPathways “Understanding the Interplay between Cultural, Biological and Subjective Factors in Drug Use Pathways” is a collaborative project supported by the European Research Area Network on Illicit Drugs (ERANID). This paper is based on independent research commissioned and funded in England by the National Institute for Health Research (NIHR) Policy Research Programme (project ref. PR-ST-0416-10001). The views expressed in this article are those of the authors and not necessarily those of the national funding agencies or ERANID. Data collection and sharing for this project was partly provided by CamCAN, funded by the UK Biotechnology and Biological Sciences Research Council (grant number BB/H008217/1), together with support from the UK Medical Research Council and University of Cambridge. The funders also had no role in study design, data collection and analysis, decision publish or preparation of the manuscript. The BRAINMINT study was supported by research grants from the Research Council of Norway (249795, 273345, 300767, 324499), the South-Eastern Norway Regional Health Authority (2014097, 2015073, 2016083, 2018076, 2019101), KG Jebsen Stiftelsen, the European Research Council under the European Union’s Horizon 2020 research and Innovation program (ERC StG Grant No. 802998), UiO:LifeScience (4MENT project), and the Department of Psychology, University of Oslo. Part of this work was performed on the TSD (Tjeneste for Sensitive Data) facilities, owned by the University of Oslo, operated and developed by the TSD service group at the University of Oslo, IT-Department (USIT), and using resources provided by UNINETT Sigma2 (#NS9666S) - the National Infrastructure for High Performance Computing and Data Storage in Norway.

## Contributions

X.L., T.J., and J.F. conceptualized the study; R.S., X.L. and T.J. designed the analytic approach; R.S. analyzed the data and visualized the results; D.A. analyzed BrainMint datasets for validation; R.S., T.J. and X.L. wrote the manuscript; S.X. helped in preprocessing the neuroimaging and genetic data; T.W.R., B.J.S. and J.F. helped in interpreting the results; D.C., Z.C. helped in visualization; J.F., T.B., G.J.B., A.H. revised the draft; L.W. was the principal investigator of BrainMint study; T.B., G.J.B., A.L.W.B., S.D., H.F., A.G., H.G., P.G., A.H., J.-L.M., M.-L.P.M., E.A., F.N., D.P.O., L.P., S.H, M.N.S., N.V., H.W., R.W., and G.S. were the principal investigators of IMAGEN Consortium; Imaging, genetic and behavioral data in the IMAGEN project were acquired and provided by the IMAGEN Consortium; All authors critically revised the manuscript. X.L., T.J., and J.F. contributed equally to this paper.

## Consortia

### IMAGEN Consortium

Tobias Banaschewski^7,8^, Gareth J. Barker^9^, Arun L.W. Bokde^10^, Sylvane Desrivières^6^, Herta Flor^11,12^, Hugh Garavan^13^, Penny Gowland^14^, Antoine Grigis^15^, Andreas Heinz^16^, Jean-Luc Martinot^17,18^, Marie-Laure Paillère Martinot^17,18,19^, Eric Artiges^17,18, 20^, Frauke Nees^7,8,21^, Dimitri Papadopoulos Orfanos^12^, Luise Poustka^22^, Michael N. Smolka^23^, Sarah Hohmann^7,8^, Nilakshi Vaidya^24^, Henrik Walter^16^, Robert Whelan^25^, Gunter Schumann^5,27^

